# Prior SARS-CoV-2 Infection and COVID-19 Vaccine Effectiveness against Outpatient Illness during Widespread Circulation of SARS-CoV-2 Omicron Variant, US Flu VE Network

**DOI:** 10.1101/2023.01.10.23284397

**Authors:** Sara Y. Tartof, Fagen Xie, Ruchi Yadav, Karen J. Wernli, Emily T. Martin, Edward A. Belongia, Manjusha Gaglani, Richard K. Zimmerman, H. Keipp Talbot, Natalie Thornburg, Brendan Flannery, Jessie R. Chung, US Flu VE Network Investigators

**Author notes:** **Corresponding author:** Jessie R. Chung, Influenza Division, Centers for Disease Control and Prevention, 1600 Clifton Rd NE, Mailstop 24/7, Atlanta, Georgia, 30329. **Alternative corresponding author/media inquiries:** Dr. Sara Y. Tartof, Kaiser Permanente Research, 100 S. Los Robles, 2^nd^ Floor, Pasadena, California, 91101. Study group team members are listed in the Acknowledgments (US Flu VE Network Investigators). **Data availability:** Data may be made available by written request to the corresponding author. **Ethics approval statement:** This activity was reviewed and approved by CDC and each site’s Institutional Review Board [See 45 C.F.R. part 46; 21 C.F.R. part 56]. **Disclosure:** The findings and conclusions are those of the authors and do not necessarily represent the official position of the Centers for Disease Control and Prevention.

## Abstract

**Background:** We estimated combined protection conferred by prior SARS-CoV-2 infection and COVID-19 vaccination against COVID-19-associated acute respiratory illness (ARI).

**Methods:** During SARS-CoV-2 Delta (B.1.617.2) and Omicron (B.1.1.529) variant circulation between October 2021 and April 2022, prospectively enrolled adult patients with outpatient ARI had respiratory and filter paper blood specimens collected for SARS-CoV-2 molecular testing and serology. Dried blood spots were tested for immunoglobulin-G antibodies against SARS-CoV-2 nucleocapsid (NP) and spike protein receptor binding domain antigen using a validated multiplex bead assay. Evidence of prior SARS-CoV-2 infection also included documented or self-reported laboratory-confirmed COVID-19. We used documented COVID-19 vaccination status to estimate vaccine effectiveness (VE) by multivariable logistic regression by prior infection status.

**Results:** 455 (29%) of 1577 participants tested positive for SARS-CoV-2 infection at enrollment; 209 (46%) case-patients and 637 (57%) test-negative patients were NP seropositive, had documented previous laboratory-confirmed COVID-19, or self-reported prior infection. Among previously uninfected patients, three-dose VE was 97% (95% confidence interval [CI], 60%– 99%) against Delta, but not statistically significant against Omicron. Among previously infected patients, three-dose VE was 57% (CI, 20%–76%) against Omicron; VE against Delta could not be estimated.

**Conclusions:** Three mRNA COVID-19 vaccine doses provided additional protection against SARS-CoV-2 Omicron variant-associated illness among previously infected participants.

## Introduction

Recovery from SARS-CoV-2 infection is followed by a period of immunologic protection against reinfection. Whether SARS-CoV-2 infection confers broad or cross-reactive protection against new SARS-CoV-2 variants is uncertain[2-4]. Vaccination and prior infection are both immunizing events, resulting in robust humoral and cellular immune response [5-9]. As such, unvaccinated individuals with prior infection could have similar levels of protection as fully vaccinated individuals with no prior infection[10-15]. Further, recent data demonstrate that combined immunity from infection and up to three doses of mRNA vaccine (i.e., hybrid immunity) surpasses that from either vaccination or infection alone[16].

Accurately identifying prior infections is difficult without prospective seroprevalence studies or laboratory-confirmed infection. Seroprevalence studies indicate that many SARS-CoV-2 infections go undetected[17, 18]. These include asymptomatic infections and those for which testing is not sought or produces false-negative results. Additionally, COVID-19 testing access and preferences vary by COVID-19 immunization status, clinical, and sociodemographic factors, likely contributing to misclassification of prior infection when relying solely on healthcare encounters data[19, 20]. Further, the rapid influx of self-administered home testing in early 2022 reduced PCR-based testing at healthcare sites[21]. Together, these factors contribute to underestimation of prior SARS-CoV-2 infection when relying on healthcare encounters data and to preferential clinical testing. Studies that have assessed protection from infection or COVID-19 vaccine effectiveness (VE) against repeat SARS-CoV-2 infection have generally relied on electronic health record (EHR) evidence of a positive laboratory test but have not considered the role of undocumented prior infection. This approach assumes the “unvaccinated” or “unexposed” individuals of the control group to be immunologically naïve. The extent of the impact of this misclassification of previously infected individuals on evaluation of protection provided by infection against new SARS-CoV-2 variants or added benefits of COVID-19 vaccination has important implications for future prevention strategies.

Previously, we estimated effectiveness of two or three doses of mRNA COVID-19 vaccines against SARS-CoV-2 Delta and Omicron variant-associated symptomatic outpatient illness[22]. Prospective collection of dried blood spots from patients provided the opportunity to assess the role of prior SARS-CoV-2 infection, including documented or serologic evidence, on VE. Here, we estimated protection conferred by hybrid immunity (prior infection and vaccination) for two- and three-dose mRNA COVID-19 vaccines via analyses stratified by prior SARS-CoV-2 infection.

## Methods

### Study Population

This study was conducted by participating institutions in the US Influenza Vaccine Effectiveness (Flu VE) Network with study sites in 7 states (CA, MI, PA, TN, TX, WA, WI) as previously described[22]. Briefly, active surveillance was conducted at outpatient healthcare facilities or COVID-19 testing sites for patients aged ≥6 months with acute illness with symptom duration of ≤10 days. Ill individuals were screened using a standard case definition for COVID-19-like illness (CLI) that included either fever, cough, or loss of taste/smell[23].

Participants completed standardized enrollment questionnaires. Data collected included participant age, gender, race, ethnicity, date of illness onset, symptoms, self-reported chronic medical condition (heart disease, lung disease, diabetes, cancer, liver/kidney disease, immune suppression, or high blood pressure), highest level of education, and high-risk exposure. High-risk exposures included healthcare work in close contact with patients, close contact in the 14 days before illness onset with a person with laboratory-confirmed SARS-CoV-2, or household member with laboratory-confirmed SARS-CoV-2 or CLI.

Participants were asked about their history of positive SARS-CoV-2 tests. Participants were asked “Prior to this illness, did you test positive for COVID-19 (by any test – e.g., rapid, PCR, or antibody test)?” Participants reporting a prior positive test were asked if the most recent positive test was <90 or ≥90 days before current illness. This activity was reviewed and approved by CDC and each site’s Institutional Review Board [See 45 C.F.R. part 46; 21 C.F.R. part 56].

### Specimen collection and laboratory testing

Participants had an oropharyngeal and nasal swab specimen collected for SARS-CoV-2 molecular testing and finger prick for collection of dried blood spots (DBS) on filter paper. Participants opting out of finger-stick blood collection were excluded from this analysis.

Preparation of DBS is described in Supplemental Methods. Briefly, research staff collected whole blood by finger stick and absorbed drops on up to 5 half-inch circles on Whatman 903 filter paper cards, which were dried at room temperature, packed with desiccant, and sent to CDC. DBS specimens were tested for immunoglobulin G (IgG) antibodies against three SARS-CoV-2 recombinant antigens representing the receptor binding domain (RBD) of the SARS-CoV-2 spike-1 protein, nucleocapsid protein (NP) and an RBD-NP hybrid antigen using a validated multiplex bead assay (FlexImmArray™ SARS-CoV-2 Human IgG Antibody Test, Tetracore, Rockville, MD) on a Luminex MAGPIX instrument with LX200 flow analyzer (Luminex Corporation, Austin, TX). Positive results for the presence of IgG antibodies against SARS-CoV-2 NP protein were defined according to the manufacturer’s instructions as median fluorescence intensity (MFI) of sample greater than 1.2-fold the MFI of the human IgG calibrator serum for NP antigen. Samples with equivocal ratios (between ≥0.9 to ≤1.2) were repeated; specimens with final ratios ≤0.9-fold that of NP antigen calibrator serum were defined as anti-NP IgG seronegative. Anti-NP IgG seropositivity was considered evidence of previous SARS-CoV-2 infection.

### Evidence of prior SARS-CoV-2 infection

Study sites searched electronic health records (EHR) for documented clinical SARS-CoV-2 tests since March 2020 for participants and extracted test type and results. Documented prior SARS-CoV-2 infection was defined as EHR documentation of positive SARS-CoV-2 RT-PCR or antigen test (including test results imported from outside the healthcare system). *Possible* prior infection was defined as a self-reported previous positive test for SARS-CoV-2 if the most recent positive test had occurred >90 days before current illness onset. *Confirmed* prior infection was defined as anti-NP seropositivity or EHR documentation of previous laboratory-confirmed SARS-CoV-2 infection >90 days before the current illness. Confirmed and possible evidence were combined for analyses; sensitivity analysis considered confirmed prior infection only.

### COVID-19 Vaccination Status

Vaccination receipt was verified in electronic immunization records as previously described[22]. All vaccine doses during the study period were monovalent mRNA products. Participants were considered vaccinated with two doses if they received two doses of either mRNA vaccine with the second administered ≥14 days before illness onset. We required at least a 16-day interval between first and second doses for Pfizer-BioNTech vaccine and at least a 23-day interval for Moderna vaccine. If illness onset occurred <7 days after a third dose of mRNA vaccine, the participant was considered to have received two doses.

Participants were considered vaccinated with three doses if they received three mRNA vaccine doses with at least a 16-day interval between second and third doses for Pfizer-BioNTech vaccine and at least a 23-day interval for Moderna vaccine with the most recent dose received ≥7 days before illness onset[24]. Those with no EHR-documented COVID-19 vaccination before illness onset were considered unvaccinated. Participants who had received one or four doses of mRNA vaccine or any non-mRNA COVID-19 vaccine were excluded (**Supplemental Figure 1**).

### Analyses

We limited all analyses to adults aged ≥18 years for whom a DBS was obtained. Characteristics were compared between those who were SARS-CoV-2-positive at enrollment versus those who tested negative. We determined the distribution of characteristics for those with 1) no evidence of prior SARS-CoV-2 infection versus those with confirmed/possible prior infection and 2) prior infection by each data source (NP serology, EHR documentation, self-report). Descriptive analyses explored the correspondence between RBD seropositivity and COVID-19 vaccination status among NP-seronegative participants.

To investigate whether COVID-19 vaccination provided additional protection beyond that conferred by prior infection, we compared odds of confirmed/possible prior infection among those with SARS-CoV-2 positive results at enrollment (i.e., cases) to those who tested negative at enrollment (i.e., controls), stratified by vaccination status (unvaccinated, two doses, three doses). Adjusted odds were estimated using logistic regression models including age, sex, race and ethnicity, site, illness onset week, self-reported chronic medical condition, and high-risk SARS-CoV-2 exposure.

VE of two or three mRNA COVID-19 vaccine doses was estimated overall, by time period/variant, and by prior infection status using the test-negative design. Cases were participants with positive SARS-CoV-2 RT-PCR results at enrollment, and controls were participants with negative SARS-CoV-2 results at enrollment. The odds of two-or three-dose mRNA COVID-19 vaccination among SARS-CoV-2-positive cases versus test-negative controls were estimated using logistic regression models adjusted for the covariates used in the model above. VE was calculated as (1 – adjusted [or unadjusted] odds ratio) x 100%. VE by number of doses received was estimated overall and by variant using time periods of predominant Delta (illness onset October 1–December 9, 2021) and Omicron circulation (illness onset December 20, 2021–April 15, 2022). These periods were selected based on the SARS-CoV-2 sequencing results from a subset of Flu VE Network SARS-CoV-2 cases[22]. Infections from December 10– 19, 2021 were excluded due to variant co-circulation. Adjusted VE was further stratified by prior infection status (possible/confirmed versus no prior infection).

## Results

From October 1, 2021 to April 15, 2022, 6,244 patients aged ≥18 years with CLI were enrolled and tested prospectively for SARS-CoV-2 infection using molecular assays. Of these, 1,883 provided DBS specimens. Of those, 1,577 met inclusion criteria for this analysis (**Table 1**).

**Table 1.**
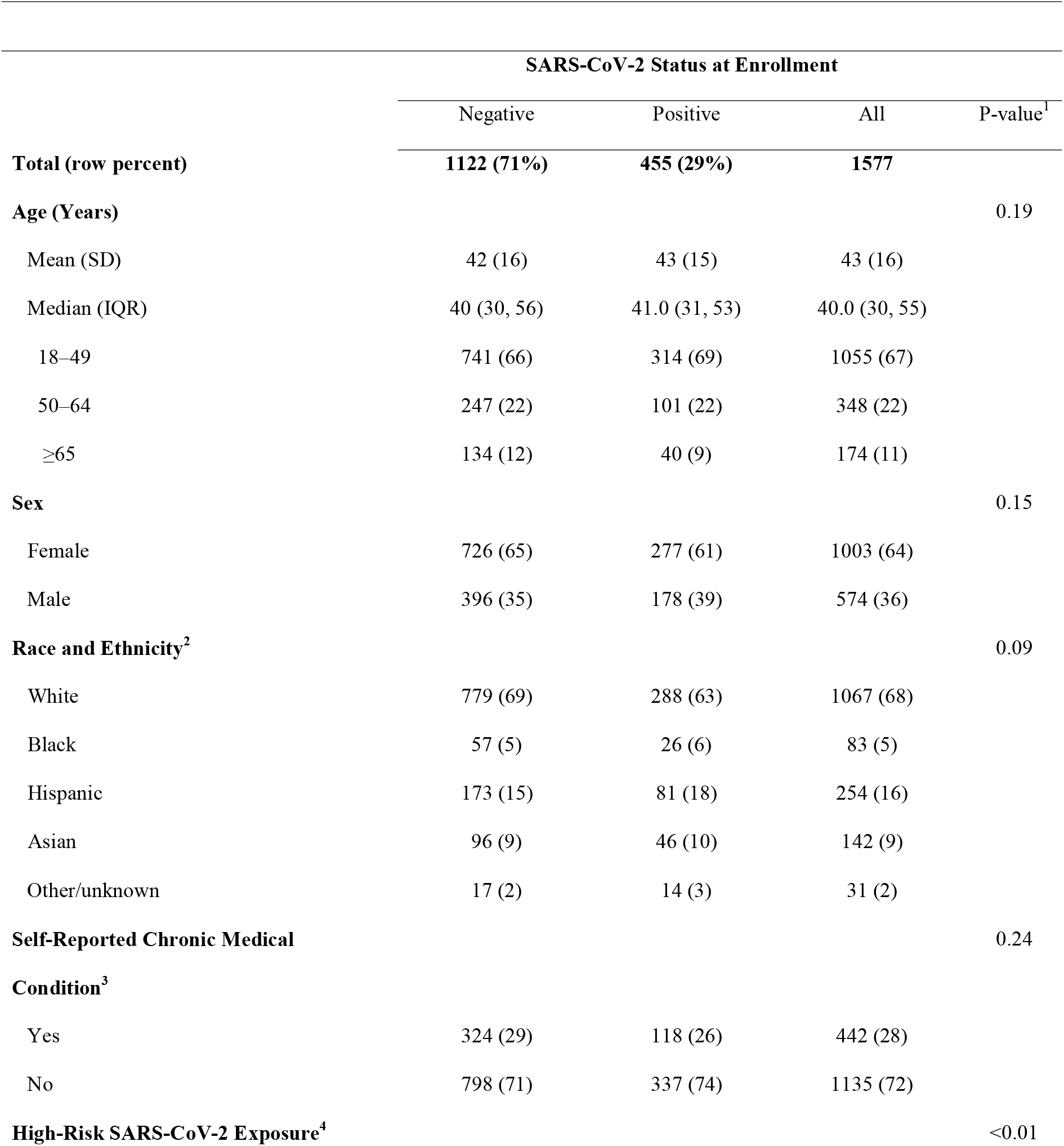
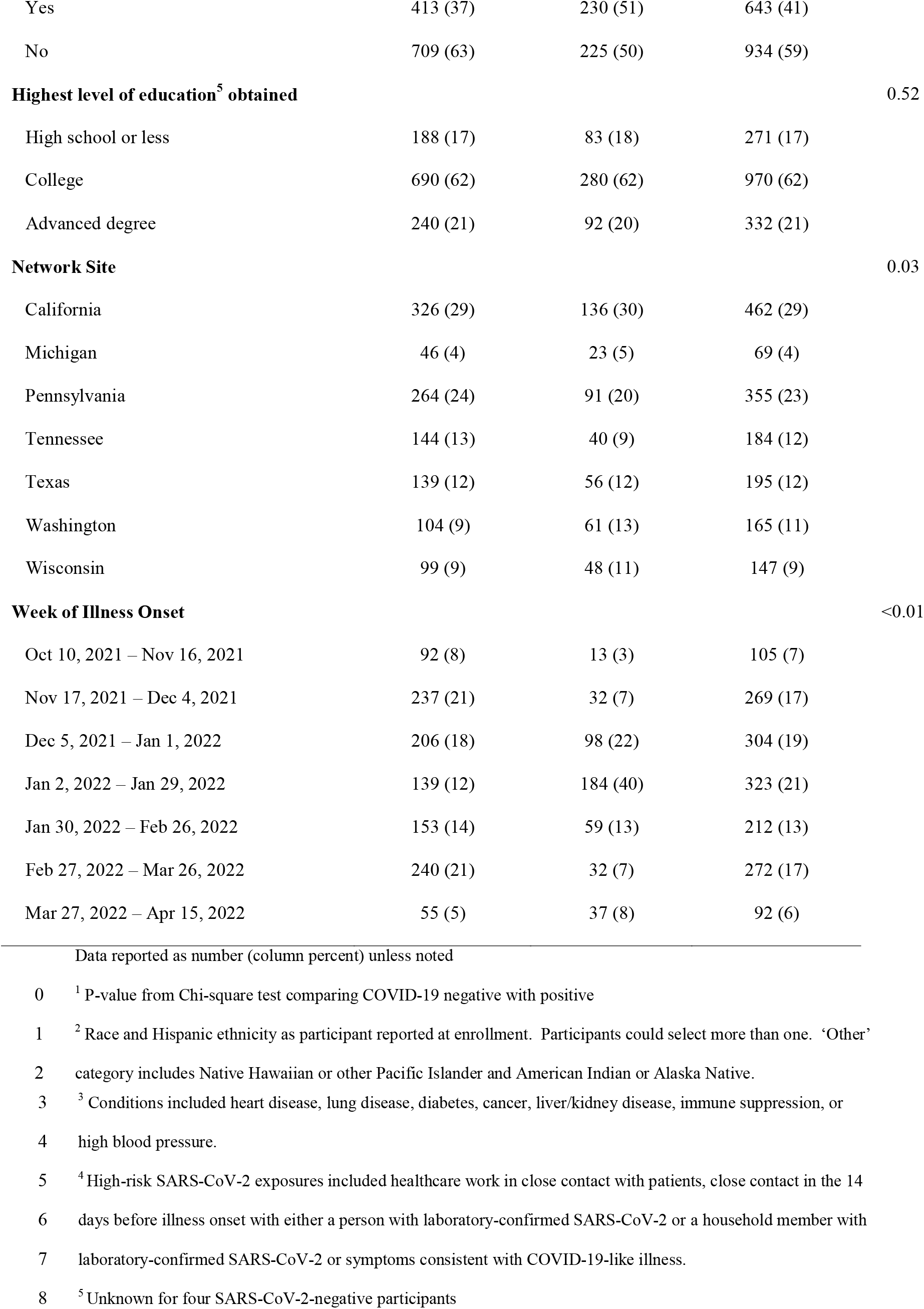
Participant characteristics by SARS-CoV-2 status at enrollment

Among these participants, mean age was 43 years (SD=16); the majority (1,033, 64%) were female, White (1,067, 68%), and college educated (970, 62%). Overall, 254 (16%) participants were Hispanic, 442 (28%) reported having a chronic medical condition, and 643 (42%) reported having a high-risk SARS-CoV-2 exposure. Of included participants, 455 (29%) tested SARS-CoV-2 positive by RT-PCR at enrollment, and 1,122 (71%) tested negative (**Table 1**). SARS-CoV-2 positivity differed significantly by high-risk exposure, study site, and week of illness onset. Median days from symptom onset to enrollment and DBS collection was three (range, 0– 13; 1 with 11 and 1 with 13).

Among 1,577 participants, 846 (54%) had evidence of confirmed/possible prior infection (**Table 2**). EHR sources of prior laboratory-confirmed SARS-CoV-2 infection documented 171 (11%) participants with prior infection. Nearly all prior infections were detected by serologic testing; 790 of 846 (93%) participants had serologic evidence of prior infection. Less than half of prior infections (241/846, 29%) were evident from more than one source. Additionally, 570 (67%) participants only had serologic evidence of prior infection, 14 (2%) only had a documented prior laboratory-confirmed SARS-CoV-2 infection, and 21 (3%) only self-reported a positive SARS-CoV-2 test that was not apparent in EHR. An additional 106 participants self-reported or had EHR documentation of an infection <90 days prior to an infection at enrollment and were excluded from analyses; 31 (29%) of these participants also had another infection >90 days prior. In all, combining three sources of prior infection data increased the proportion of participants classified as having prior SARS-CoV-2 infection by nearly 5-fold compared to using EHR documentation alone to define prior infection status.

**Table 2.** Characteristics of participants with prior SARS-CoV-2 infection category

|  |  |  |  |  |  | Prior SARS-CoV-2 infection status |  |  |  |  |  |
| --- | --- | --- | --- | --- | --- | --- | --- | --- | --- | --- | --- |
|  | N | No evidence of prior infection |  | Confirmed<sup>†,‡</sup> or possible<sup>§</sup> prior infection |  | Serologic<sup>†</sup> |  | Documented<sup>‡</sup> |  | Self-reported<sup>§</sup> |  |
|  | N | No. | % | No. | % | No. | %<sup>¶</sup> | No. | %<sup>¶</sup> | No. | %<sup>¶</sup> |
|  | 1577 | 731 | 46 | 846 | 54 | 790 | 50 | 171 | 11 | 212 | 13 |
| **Age (years)** |  |  |  |  |  |  |  |  |  |  |  |
| Mean (SD) |  | 42.5 (15.3) |  | 43.1 (15.9) |  | 43.3 (16.0) |  | 43.2 (14.6) |  | 39.3 (14.7) |  |
| 18-49 | 1055 | 506 | 48 | 549 | 52 | 507 | 48 | 111 | 11 | 157 | 15 |
| 50-64 | 348 | 150 | 43 | 198 | 57 | 189 | 54 | 45 | 13 | 40 | 11 |
| ≥65 | 174 | 75 | 43 | 99 | 57 | 94 | 54 | 15 | 9 | 15 | 9 |
| **Sex** |  |  |  |  |  |  |  |  |  |  |  |
| Female | 1003 | 458 | 46 | 545 | 54 | 509 | 51 | 125 | 12 | 148 | 15 |
| Male | 574 | 273 | 48 | 301 | 52 | 281 | 49 | 46 | 8 | 64 | 11 |
| **Race and Ethnicity<sup>††</sup>** |  |  |  |  |  |  |  |  |  |  |  |
| White | 1067 | 482 | 45 | 585 | 55 | 540 | 51 | 115 | 11 | 149 | 14 |
| Black | 83 | 39 | 47 | 44 | 53 | 44 | 53 | 4 | 5 | 11 | 13 |
| Hispanic | 254 | 112 | 44 | 142 | 56 | 135 | 53 | 39 | 15 | 37 | 15 |
| Asian | 142 | 81 | 57 | 61 | 43 | 57 | 40 | 9 | 6 | 11 | 8 |
| Other/Unknown | 31 | 17 | 55 | 14 | 45 | 14 | 45 | 4 | 13 | 4 | 13 |
| **Highest level of education obtained<sup>††</sup>** |  |  |  |  |  |  |  |  |  |  |  |
| High School or less | 271 | 123 | 45 | 148 | 55 | 137 | 51 | 34 | 13 | 40 | 15 |
| College | 970 | 441 | 45 | 529 | 55 | 494 | 51 | 112 | 12 | 151 | 16 |
| Advanced Degree | 332 | 166 | 50 | 166 | 50 | 156 | 47 | 24 | 7 | 21 | 6 |
| **Self-reported Chronic Medical Condition<sup>§§</sup>** |  |  |  |  |  |  |  |  |  |  |  |
| Yes | 442 | 213 | 48 | 229 | 52 | 212 | 48 | 45 | 10 | 53 | 12 |
| No | 1135 | 518 | 46 | 617 | 54 | 578 | 51 | 126 | 11 | 159 | 14 |
| **High-Risk SARS-CoV-2 exposure<sup>¶¶</sup>** |  |  |  |  |  |  |  |  |  |  |  |
| Yes | 643 | 281 | 44 | 362 | 56 | 340 | 53 | 76 | 12 | 103 | 16 |
| No | 934 | 450 | 48 | 484 | 52 | 450 | 48 | 95 | 10 | 109 | 12 |
| **Number of COVID-19 vaccine doses<sup>+++</sup>** |  |  |  |  |  |  |  |  |  |  |  |
| Unvaccinated | 289 | 135 | 47 | 154 | 53 | 137 | 47 | 43 | 15 | 63 | 22 |
| 2-dose | 647 | 389 | 60 | 258 | 40 | 235 | 36 | 70 | 11 | 81 | 13 |
| 3-dose | 641 | 207 | 32 | 434 | 68 | 418 | 65 | 58 | 9 | 68 | 11 |
| **SARS-CoV-2 test result at enrollment** |  |  |  |  |  |  |  |  |  |  |  |
| Positive | 455 | 246 | 54 | 209 | 46 | 192 | 42 | 38 | 8 | 41 | 9 |
| Negative | 1122 | 485 | 43 | 637 | 57 | 598 | 53 | 133 | 12 | 171 | 15 |

<sup>†</sup> Nucleocapsid protein (NP) seropositive by multiplex SARS-CoV-2 bead assay (FlexImmArray, Tetracore).
<sup>‡</sup> Laboratory confirmed SARS-CoV-2 infection documented in electronic health record (EHR) >90 days before onset of current illness.
<sup>§</sup> Participant self-report of prior confirmed COVID-19 or positive SARS-CoV-2 test >90 days before onset of current illness
<sup>¶</sup> Denominator is the row total for the header row. Categories of prior SARS-CoV-2 infection status are not mutually exclusive and sum to more than 100%.
<sup>††</sup> Race and Hispanic ethnicity as participant reported at enrollment. Participants could select more than one. ‘Other’ category includes Native Hawaiian or other Pacific Islander and American Indian or Alaska Native.
<sup>‡‡</sup> Unknown for four participants.
<sup>§§</sup> Conditions included heart disease, lung disease, diabetes, cancer, liver/kidney disease, immune suppression, or high blood pressure
<sup>¶¶</sup> High-risk SARS-CoV-2 exposures included healthcare work in close contact with patients, close contact in the 14 days before illness onset with either a person with laboratory-confirmed SARS-CoV-2 or a household member with laboratory-confirmed SARS-CoV-2 or symptoms consistent with COVID-19-like illness.
<sup>†††</sup> Number of COVID-19 vaccine doses reported in a participant’s electronic health record.

Among participants included in this analysis, 641 (41%) had received three doses and 647 (41%) had received two doses of monovalent mRNA vaccines ≥14 days before illness onset; 289 (18%) were unvaccinated at the time of enrollment. Presence of antibodies to SARS-CoV-2 spike protein RBD antigen correlated with EHR-documented COVID-19 vaccination status (**Table 3**). However, 18 of 647 participants with two-doses (3%) and 9 of 641 participants with three-doses (1%) were RBD-antibody seronegative at enrollment. Of these 27 RBD-seronegative patients with prior vaccination, the mean and median time since second dose (n=18) was 240 and 235 days, respectively; mean and median time since third dose was 83 and 78 days, respectively (n=18). About half (48%) of the 27 RBD-seronegative vaccinated participants had chronic medical conditions, compared to 29% of seropositive vaccinated participants.

**Table 3.** Seroprevalence of antibodies against SARS-CoV-2 Spike protein Receptor Binding Domain (RBD) by COVID-19 vaccination status and evidence of prior SARS-CoV-2 infection^†^

|  | **Total<sup>†</sup>** | **No. (column %)**<br>**Seropositive<sup>†</sup>** | **No. (column %)**<br>**Seronegative** | **P-value<sup>§</sup>** |
| --- | --- | --- | --- | --- |
| **Number of COVID-19 Vaccine Doses<sup>¶</sup>** |  |  |  | <0.01 |
| Unvaccinated | 267 | 172 (13) | 95 (78) |  |
| 2 doses | 613 | 595 (45) | 18 (15) |  |
| 3 doses | 561 | 552 (42) | 9 (7) |  |
| **Confirmed or possible prior SARS-CoV-2 infection<sup>††</sup>** |  |  |  | <0.01 |
| Unvaccinated | 148 | 138 (17) | 10 (71) |  |
| 2 doses | 252 | 251 (31) | 1 (7) |  |
| 3 doses | 429 | 426 (52) | 3 (21) |  |
| **No evidence of prior SARS-CoV-2 infection<sup>††</sup>** |  |  |  | <0.01 |
| Unvaccinated | 119 | 34 (7) | 85 (79) |  |
| 2 doses | 361 | 344 (68) | 17 (16) |  |
| 3 doses | 132 | 126 (25) | 6 (6) |  |

RBD, SARS-CoV-2 protein receptor binding domain
<sup>†</sup> Seropositive for SARS-CoV-2 Spike Protein RBD.
<sup>‡</sup> Excludes 136 participants with indeterminate serostatus for RBD (n=7) or NP (n=129) antibody. 22 participants with indeterminate serostatus were unvaccinated, 34 received 2 doses of vaccine, and 80 received 3 doses of vaccine.
<sup>§</sup> P-value for Chi-square test comparing proportion seropositive to seronegative.
<sup>¶</sup> Vaccination status based on number of doses documented in electronic medical record received $\geq 14$ days before illness onset for 2<sup>nd</sup> dose or $\geq 7$ days before illness onset for 3<sup>rd</sup> dose.
<sup>††</sup> Includes seropositive nucleocapsid protein (NP) antibody assay, electronic medical record documented positive SARS-CoV-2 molecular or antigen test, or participant self-report of positive SARS-CoV-2 positive laboratory test >90 days before onset of current illness.
<sup>‡‡</sup> Participants NP antibody seronegative without EHR-documented or self-reported positive COVID-19 test >90 days before onset of current illness

Among 148 unvaccinated participants with confirmed/possible prior SARS-CoV-2 infection, 138 (93%) were RBD-antibody seropositive and NP seronegative, indicating presence of anti-Spike protein antibody from prior infection, undocumented vaccination, or both. In contrast, 34 (29%) of 119 unvaccinated participants without confirmed/possible prior SARS-CoV-2 infection were RBD-antibody seropositive and NP seronegative at enrollment, suggesting serologic evidence of prior infection or undocumented vaccination.

### Prior SARS-CoV-2 infection and risk of SARS-CoV-2 at enrollment

The adjusted odds of having SARS-CoV-2 at enrollment tended to be reduced for those with prior infection, regardless of vaccination status (**Table 4)**. For the entire enrollment period, prior SARS-CoV-2 infection was associated with reduced odds of testing positive for SARS-CoV-2 among participants who had received two COVID-19 mRNA doses (aOR=0.47; 95% CI, 0.30– 0.76) or three doses (aOR=0.76; 95% CI, 0.49–1.19), but confidence intervals were wide and not statistically significant for three doses. Among two-dose vaccine recipients during the Omicron-variant period, the odds of testing positive were significantly reduced among participants with prior infection vs. those without prior infection (OR=0.39, 95% CI, 0.22–0.70). Similarly, the odds of testing positive among three-dose recipients was 0.81 (95% CI, 0.51–1.29) for those with prior infection vs. those without prior infection. Findings were similar when only EHR documentation was used to define prior infection status (**sTable 1**).

**Table 4.** Odds Ratios for SARS-CoV-2 infection among 1,577 ill participants with and without evidence^†^ of prior SARS-CoV-2 infection by COVID-19 vaccination status^‡^ and SARS-CoV-2 virus type.

|  |  |  |  |  | Unadjusted OR<br>[95% CI] | Adjusted<sup>§</sup> OR<br>[95% CI] |
| --- | --- | --- | --- | --- | --- | --- |
|  | Prior infection | Total | No. SARS-CoV-2 positive | % positive |  |  |
| **Overall** |  |  |  |  |  |  |
| Unvaccinated | No Prior infection | 135 | 60 | 44 | Referent |  |
|  | Prior infection | 154 | 45 | 29 | 0.52 [0.32, 0.84] | 0.59 [0.32, 1.08] |
| 2 doses | No Prior infection | 389 | 129 | 33 | Referent |  |
|  | Prior infection | 258 | 57 | 22 | 0.57 [0.40, 0.82] | 0.47 [0.30, 0.76] |
| 3 doses | No Prior infection | 207 | 57 | 28 | Referent |  |
|  | Prior infection | 434 | 107 | 25 | 0.86 [0.59, 1.25] | 0.76 [0.49, 1.19] |
| **Delta variant** |  |  |  |  |  |  |
| Unvaccinated | No Prior infection | 43 | 14 | 33 | Referent |  |
|  | Prior infection | 37 | 5 | 14 | 0.32 [0.10, 1.0] | 0.41 [0.11, 1.52] |
| 2 doses | No Prior infection | 189 | 20 | 11 | Referent |  |
|  | Prior infection | 83 | 8 | 10 | 0.90 [0.38, 2.14] | 0.84 [0.32, 2.22] |
| 3 doses | No Prior infection | 31 | 1 | 3 | Referent |  |
|  | Prior infection | 33 | 1 | 3 | 0.75 [0.05, 12.48] | 0.58 [0.03, 12.56] |
| **Omicron variant** |  |  |  |  |  |  |
| Unvaccinated | No Prior infection | 74 | 38 | 51 | Referent |  |
|  | Prior infection | 109 | 38 | 35 | 0.51 [0.40, 0.82] | 0.86 [0.37, 2.01] |
| 2 doses | No Prior infection | 157 | 93 | 59 | Referent |  |
|  | Prior infection | 158 | 47 | 30 | 0.29 [0.18, 0.46] | 0.39 [0.22, 0.70] |
| 3 doses | No Prior infection | 162 | 53 | 33 | Referent |  |
|  | Prior infection | 371 | 104 | 28 | 0.80 [0.54, 1.19] | 0.81 [0.51, 1.29] |

CI, confidence interval; OR, odds ratio
<sup>†</sup> Includes seropositive nucleocapsid protein (NP) antibody assay, electronic medical record documented positive SARS-CoV-2 molecular or antigen test, or participant self-report of positive SARS-CoV-2 positive laboratory test >90 days before onset of current illness.
<sup>‡</sup> Vaccination status based on number of doses documented in electronic medical record received $\geq 14$ days before illness onset for 2<sup>nd</sup> dose or $\geq 7$ days before illness onset for 3<sup>rd</sup> dose.
<sup>§</sup> Model adjusted for age, sex, race/ethnicity, site, illness onset week, self-reported chronic medical condition, high-risk SARS-CoV-2 exposure.

### Prior SARS-CoV-2 infection and COVID-19 vaccine effectiveness

Among 455 patients who tested SARS-CoV-2 positive at enrollment, 186 (41%) had received two COVID-19 mRNA vaccine doses, 164 (36%) had received three doses, and 209 (46%) had confirmed/possible evidence of prior SARS-CoV-2 infection. Among 1,122 SARS-CoV-2-negative participants, 461 (41%) and 477 (43%) had received two or three mRNA vaccine doses, respectively, and 637 (57%) had evidence of prior SARS-CoV-2 infection.

Overall, adjusted VE for two COVID-19 vaccine doses among patients with no evidence of prior SARS-CoV-2 infection tended to be lower than VE estimates among patients with confirmed/possible prior infection, although neither estimate was statistically significant (**Table 5**). During the SARS-CoV-2 Omicron variant-predominant period, two-dose VE was 45% (95% CI, -4 to 70) among those with confirmed/possible prior infection versus -53% (95% CI, -205 to 23) among those without prior infection; three-dose VE was 57% (95% CI, 20 to 76) versus 23% (95% CI, -72 to 65). Fewer breakthrough infections during the Delta-variant-predominant period did not allow for meaningful comparisons between strata. Findings were similar when only EHR documentation was used to define prior infection status (**sTable 2**).

**Table 5.** Adjusted^†^ vaccine effectiveness against COVID-19 by SARS-CoV-2 variant stratified by prior infection status^‡^

|  | Vaccination<sup>§</sup> | Total | SARS-CoV-2 positive | % positive | Unadjusted VE<br>[95% CI] | Adjusted<sup>†</sup> VE<br>[95% CI] |
| --- | --- | --- | --- | --- | --- | --- |
| **Overall** |  |  |  |  |  |  |
| No prior infection | Unvaccinated | 135 | 60 | 44 | Referent |  |
|  | 2 doses | 389 | 129 | 33 | 38 [8, 58] | 17 [-40, 51] |
|  | 3 doses | 207 | 57 | 28 | 53 [25, 70] | 53 [10, 76] |
| With prior infection | Unvaccinated | 154 | 45 | 29 | Referent |  |
|  | 2 doses | 258 | 57 | 22 | 31 [-8, 56] | 42 [-1, 66] |
|  | 3 doses | 434 | 107 | 25 | 21 [-19, 47] | 55 [21, 74] |
| **Delta variant** |  |  |  |  |  |  |
| No prior infection | Unvaccinated | 43 | 14 | 33 | Referent |  |
|  | 2 doses | 189 | 20 | 11 | 76 [46, 89] | 70 [11, 90] |
|  | 3 doses | 31 | 1 | 3 | 93 [44, 99] | 97 [60, 100] |
| With prior infection | Unvaccinated | 37 | 5 | 14 | Referent |  |
|  | 2 doses | 83 | 8 | 10 | 32 [-125, 79] | 23 [-225, 82] |
|  | 3 doses | 33 | 1 | 3 | 84 [-44, 98] | 87 [-86, 99] |

| Omicron variant |  |  |  |  |  |  |
| --- | --- | --- | --- | --- | --- | --- |
| No prior infection | Unvaccinated | 74 | 38 | 51 | Referent |  |
|  | 2 doses | 157 | 93 | 59 | -38 [-140, 21] | -53 [-205, 23] |
|  | 3 doses | 162 | 53 | 33 | 54 [19, 74] | 23 [-72, 65] |
| With prior infection | Unvaccinated | 109 | 38 | 35 | Referent |  |
|  | 2 doses | 158 | 47 | 30 | 21 [-33, 53] | 45 [-4, 70] |
|  | 3 doses | 371 | 104 | 28 | 27 [-15, 54] | 57 [20, 76] |

CI, confidence interval; VE, vaccine effectiveness
<sup>†</sup> Models adjusted for age, sex, race/ethnicity, site, illness onset week, self-reported chronic medical condition, high-risk SARS-CoV-2 exposure.
<sup>‡</sup> Includes seropositive nucleocapsid protein (NP) antibody assay, electronic medical record documented positive SARS-CoV-2 molecular or antigen test, or participant self-report of positive SARS-CoV-2 positive laboratory test >90 days before onset of current illness.
<sup>§</sup> Vaccination status based on number of doses documented in electronic medical record received $\geq 14$ days before illness onset for 2<sup>nd</sup> dose or $\geq 7$ days before illness onset for 3<sup>rd</sup> dose.

## Discussion

Understanding the magnitude of protection from infection and the potential additional protection from COVID-19 vaccination (i.e., ‘hybrid immunity’) against illness associated with new SARS-CoV-2 variant viruses is important for future COVID-19 vaccine programs. Here, we found that confirmed or possible prior infection was associated with protection against current SARS-CoV-2 infection, regardless of vaccination status during the early 2022 Omicron-variant wave. Vaccination-conferred immunity among patients with no evidence of prior SARS-CoV-2 infection appeared to vary by variant, where two and three doses of COVID-19 mRNA vaccines provided significant protection against SARS-CoV-2-associated illness during the Delta variant-predominant period, but no significant protection during the Omicron-variant period.

For patients with evidence of prior infection, three doses of COVID-19 mRNA monovalent vaccines provided additional protection against illness during the Omicron-variant period, but we could not measure statistically significant protection from two vaccine doses or among previously uninfected participants who received two or three vaccine doses. Cases with prior infection were insufficient in the Delta period to assess the role of vaccine during that time. Protection from primary-series vaccination is short-lived against Omicron infections[25, 26], and vaccination with monovalent formulations of mRNA vaccines have been demonstrated to confer weaker protection against Omicron compared with Delta.[27] These findings are also consistent with work demonstrating that a third dose of BNT162b2 vaccine was needed for induction of consistent neutralizing antibody titers against either BA.1 or BA.2.[28, 29] Relatively weaker naturally acquired protective immunity against Omicron in our findings is consistent with waning over time and/or immune evasion of new variants in the absence of boosting with updated vaccines.

A matched test-negative study from Qatar also found that three vaccine doses and previous infection conferred the greatest protection against SARS-CoV-2 Omicron-variant infection, compared with two doses and prior infection and three doses without prior infection[16]. However, those with two doses of vaccine and previous infection were afforded similar levels of protection as those with three doses and no previous infection, which was similar to protection from previous infection alone. In our study, while estimates for three doses and no previous infection and those with two doses and prior infection are not meaningfully different, sample size limitations preclude definitive comparison. Further, our US-based population has very high levels of immunity by either vaccination or infection, which may also lessen our ability to discriminate between levels of immunity.

Most persons with SARS-CoV-2 infection generate detectable anti-SARS-CoV-2 antibodies, with studies reporting seroconversion rates >90%[30, 31]. The consistency in antibody response allows for highly sensitive serologically based detection methods for prior infections. Here, we found that EHR determination of prior infection alone was not very informative; supplementing those data with NP serology resulted in a 5-fold increase in evidence of prior infection. While EHR-documented evidence of prior infection is highly specific, it is not sensitive and likely misses a large proportion of those not tested, tested outside the home healthcare system, or self-tested with antigen-based kits[21]. Yet, most large-scale VE studies to date define prior infection by EHR data sources alone. While NP serology has higher sensitivity and specificity to estimate prior infection than other modes of ascertainment, future data regarding sensitivity and specificity by time since infection are needed.

RBD serology findings can also inform our understanding of serologic test performance. Seropositivity for SARS-CoV-2 spike protein RBD among unvaccinated patients likely indicates prior infection. However, in this study, we identified 34 patients with RBD seropositivity who were seronegative for NP antibodies and were also unvaccinated, suggesting that some prior infections, including potentially asymptomatic infections, may have been missed due to waning of anti-NP antibody, which occurs more quickly than RBD antibodies[32]. It is also possible that these patients had prior COVID-19 vaccination events that were not captured in the EHR. Conversely, not all vaccinated people were RBD seropositive, where 27 individuals were RBD-seronegative but had documentation of two or three doses of mRNA vaccine. As with NP serology, further studies are needed on infection- and vaccination-elicited RBD antibody durability and performance as a biomarker in epidemiologic research.

There are limitations to this study. For NP serology findings, timing of infection and infecting variant are unknown; we required that self-reported and EHR-documented infections occurred ≥90 days prior to illness onset, but we were not able to gauge time of infection using serologic evidence. It is possible that an infection detected at the time of enrollment could have been a prolonged acute respiratory illness rather than a “previous infection” as defined by NP serology. Some vaccinated persons may not seroconvert for NP after infection[33], which could lead to misclassification of prior infection in a subset of participants. Our sample size was limited, and some estimates were imprecise, which constrained our ability to compare trends in hybrid immunity over time and variant. Further, variant-specific analyses were defined by secular period rather than sequencing, which may result in some misclassification. We were not able to differentiate BA.1 vs. BA.2. However, studies have demonstrated neutralizing antibody titers against BA.1 and BA.2 were similar, and that robust neutralizing antibody titers against BA.2 developed in those previously infected with BA.1, which suggests a substantial degree of cross-reactive immunity[28]. We only capture those that sought medical care or testing in an outpatient healthcare setting. While those that seek RT-PCR testing may differ from those who test at home or do not test, the test-negative design equalizes healthcare seeking behavior between our comparison groups and minimizes potential bias by vaccination status. Finally, our analyses are based on the monovalent mRNA COVID-19 vaccine formulations and may not correspond to findings derived from bivalent formulations.

Improved understanding of cross-protection elicited by infection and vaccination is needed to inform future vaccine formulations and vaccination recommendations. The optimal vaccination strategy for previously infected individuals would boost protective immunity from natural infection. If natural immunity fosters cross-protection against emerging variants, formulations that include more cross-reactive antigens may be necessary to improve VE. As new variants continue to emerge, ongoing analyses of cross-protection between strains will be important to inform vaccine programs.

## Data Availability

All data produced in the present study are available upon reasonable request to the corresponding author.

## Acknowledgments

US Flu VE Network Investigators: Kaiser Permanente Southern California: Vennis Hong, Ana Florea, Jen Ku, Jeniffer Kim, Sally Shaw, Bruno Lewin, Michael Aragones; Kaiser Permanente Washington Health Research Institute: Erika Kiniry C. Hallie Phillips, Stacie Wellwood; University of Michigan: Arnold S. Monto, Caroline Cheng; Henry Ford Medical Center: Lois Lamerato; Marshfield Clinic Research Institute: Huong Q. McLean, Jennifer P. King, Jennifer K. Meece; Baylor Scott & White Health: Michael E. Smith, Kayan Dunnigan, Eric Hoffman; University of Pittsburgh: Krissy Moehling Geffel, Louise Taylor, Mary Patricia Nowalk; Vanderbilt University: Carlos G. Grijalva, Yuwei Zhu, James D. Chappelle; US Centers for Disease Control and Prevention: Eric Rogier, Venkatachalam Udhayakumar, Devyani Joshi, Sara S. Kim, Jessie R. Chung, Manish Patel.

**Supplemental Methods**. FlexImmArray SARS-CoV-2 IgG Assay kit

**Supplemental Figure**. Study flow diagram

**Supplemental Table 1**. Odds Ratios for SARS-CoV-2 infection among ill participants with and without evidence of prior SARS-CoV-2 infection by COVID-19 vaccination status and SARS-CoV-2 virus type.

**Supplemental Table 2**. Adjusted vaccine effectiveness against COVID-19 by SARS-CoV-2 variant stratified by prior infection status

