## Supplementary figures and images for "Prior SARS-CoV-2 Infection and COVID-19 Vaccine Effectiveness against Outpatient Illness during Widespread Circulation of SARS-CoV-2 Omicron Variant, US Flu VE Network"

### Supplemental Figure

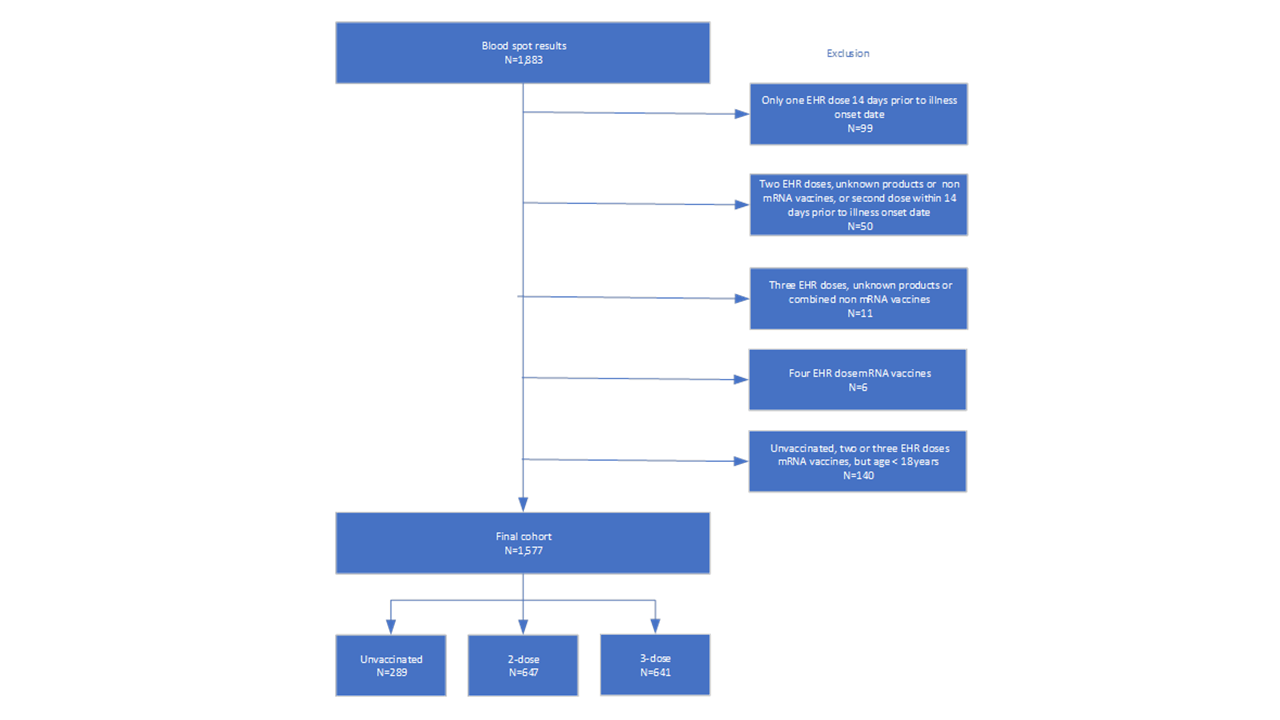
