## Supplemental Methods for "Prior SARS-CoV-2 Infection and COVID-19 Vaccine Effectiveness against Outpatient Illness during Widespread Circulation of SARS-CoV-2 Omicron Variant, US Flu VE Network"

sMethods. *FlexImmArray SARS-CoV-2 IgG Assay kit:*

This assay utilizes magnetic microspheres coupled with unique recombinant proteins specific for SARS-CoV-2 (RBD, NP, and RBD/NP fusion). The assay also includes four different internal controls for monitoring each step of assay performance. External positive control, negative control, and calibrator reagents are also provided in the kit and run-in duplicate on every assay plate. Extracted samples from dry blood spots tested in duplicate were diluted 1:400 in assay kit sample dilution buffer, immediately mixed with the antigen-coated microspheres in a 96-well plate and incubated for 20 minutes by gentle shaking at room temperature protected from light. Plates were washed four times with assay wash buffer, and DBS elute IgG antibodies were detected using anti-human IgG conjugated to phycoerythrin by incubation at 20 min under gentle shaking protected from light. The microspheres resuspended in wash buffer were analyzed using a Luminex MAGPIX instrument and a Luminex LX 200 flow analyzer (Luminex Corporation, Austin, TX) with a target of 50 beads per region. The assay cutoff established by the manufacturer were used for scoring test positive samples and any indeterminate (equivocal) samples were repeated as recommended by the test manufacturer.
