## Supplemental Table 1 and Table 2 for "Prior SARS-CoV-2 Infection and COVID-19 Vaccine Effectiveness against Outpatient Illness during Widespread Circulation of SARS-CoV-2 Omicron Variant, US Flu VE Network"

**Supplemental Table 1.** Odds Ratios for SARS-CoV-2 infection among ill participants with and without evidence^1^ of prior SARS-CoV-2 infection by COVID-19 vaccination status^2^ and SARS-CoV-2 virus type.

|  | **Prior infection** | **Total** | **SARS-CoV-2 positive** | **% positive** | **Unadjusted OR**  **[95% CI]** | **Adjusted^3^ OR**  **[95% CI]** |
| --- | --- | --- | --- | --- | --- | --- |
| **Overall** |  |  |  |  |  |  |
| Unvaccinated | No Prior infection | 246 | 92 | 37 | Referent |  |
|  | Prior infection | 43 | 13 | 30 | 0.73 [0.36, 1.46] | 0.67 [0.27, 1.65] |
| 2 doses | No Prior infection | 577 | 174 | 30 | Referent |  |
|  | Prior infection | 70 | 12 | 17 | 0.48 [0.25, 0.91] | 0.45 [0.21, 0.95] |
| 3 doses | No Prior infection | 583 | 151 | 26 | Referent |  |
|  | Prior infection | 58 | 13 | 22 | 0.83 [0.43, 1.57] | 0.68 [0.31, 1.47] |
| **Omicron variant** |  |  |  |  |  |  |
| Unvaccinated | No Prior infection | 146 | 63 | 43 | Referent |  |
|  | Prior infection | 37 | 13 | 35 | 0.71 [0.34, 1.51] | 0.94 [0.32, 2.70] |
| 2 doses | No Prior infection | 275 | 128 | 47 | Referent |  |
|  | Prior infection | 40 | 12 | 30 | 0.49 [0.24, 1.01] | 0.54 [0.23, 1.28] |
| 3 doses | No Prior infection | 479 | 144 | 30 | Referent |  |
|  | Prior infection | 54 | 13 | 24 | 0.74 [0.38, 1.42] | 0.70 [0.32, 1.52] |

CI, confidence interval; OR, odds ratio

^1^ Only includes electronic medical record documented positive SARS-CoV-2 molecular or antigen test >90 days before onset of current illness.

^2^ Vaccination status based on number of doses documented in electronic medical record received ≥14 days before illness onset for 2^nd^ dose or ≥7 days before illness onset for 3^rd^ dose.

^3^ Model adjusted for age, sex, race/ethnicity, site, illness onset week, self-reported chronic medical condition, high-risk SARS-CoV-2 exposure.

**Supplemental Table 2.** Adjusted^1^ vaccine effectiveness against COVID-19 by SARS-CoV-2 variant stratified by prior infection status^2^

|  | **Vaccination^3^** | **Total** | **SARS-CoV-2 positive** | **% positive** | **Unadjusted VE**  **[95% CI]** | **Adjusted^1^ VE**  **[95% CI]** |
| --- | --- | --- | --- | --- | --- | --- |
| **Overall** |  |  |  |  |  |  |
| No prior infection | Unvaccinated | 246 | 92 | 37 | Referent |  |
|  | 2 doses | 577 | 174 | 30 | 28 [1, 47] | 22 [-13, 47] |
|  | 3 doses | 583 | 151 | 26 | 42 [20, 57] | 57 [35, 72] |
| With prior infection | Unvaccinated | 43 | 13 | 30 | Referent |  |
|  | 2 doses | 70 | 12 | 17 | 52 [-17, 81] | 76 [10, 94] |
|  | 3 doses | 58 | 13 | 22 | 33 [-63, 73] | 80 [2, 96] |
| **Omicron variant** |  |  |  |  |  |  |
| No prior infection | Unvaccinated | 146 | 63 | 43 | Referent |  |
|  | 2 doses | 275 | 128 | 47 | -15 [-92, 23] | -5 [-68, 34] |
|  | 3 doses | 479 | 144 | 30 | 43 [17, 61] | 44 [-8, 66] |
| With prior infection | Unvaccinated | 37 | 13 | 35 | Referent |  |
|  | 2 doses | 40 | 12 | 30 | 21 [-106, 70] | 68 [-23, 92] |
|  | 3 doses | 54 | 13 | 24 | 42 [-47, 77] | 75 [-35, 95] |

CI, confidence interval; VE, vaccine effectiveness

^1^ Models adjusted for age, sex, race/ethnicity, site, illness onset week, self-reported chronic medical condition, high-risk SARS-CoV-2 exposure.

^2^ Only includes electronic medical record documented positive SARS-CoV-2 molecular or antigen test >90 days before onset of current illness.

^3^ Vaccination status based on number of doses documented in electronic medical record received ≥14 days before illness onset for 2^nd^ dose or ≥7 days before illness onset for 3^rd^ dose.
